# Effectiveness of an over-the-counter self-fitting hearing aid compared to an audiologist-fitted hearing aid: A randomized clinical trial

**DOI:** 10.1101/2022.12.12.22283331

**Authors:** Karina C. De Sousa, Vinaya Manchaiah, David R. Moore, Marien A. Graham, De Wet Swanepoel

## Abstract

**Importance:** Hearing loss is a highly prevalent condition, with numerous debilitating consequences when left untreated. However, less than 20% of adults with hearing loss in the United States use hearing aids. Over-the-counter (OTC) hearing aids became available in October 2022 to improve access and affordability. However, clinical effectiveness studies of available OTC hearing aids using the existing devices in the market are limited.

**Objective:** To compare the clinical effectiveness of a self-fitting OTC hearing aid with remote support to a hearing aid fitted using audiologist best practices.

**Design, setting, and participants:** A randomized parallel-assigned clinical effectiveness trial was conducted between April to August 2022. Sixty-eight adults with self-perceived mild-to-moderate hearing loss were recruited and randomly assigned to either the self-fitting (SF) group or an audiologist-fitted (AF) group. Following hearing aid fitting, participants first completed a two-week, take-home field trial without any support. Access to fine-tuning for both groups was only available after the two-week trial. The level of support and adjustment was done remotely for the SF group per request and by the audiologist for the AF group. Participants were then re-assessed after an additional four-week take-home trial.

**Interventions:** A commercially available self-fitting OTC hearing aid was provided to participants in the SF group who were expected to set up the hearing aids using the commercially supplied instructional material and accompanying smartphone app. In the AF group, audiologists fitted the same hearing aid according to a commonly used (NAL-NL2) prescriptive gain target using real-ear verification with hearing aid use instruction.

**Main Outcomes and Measures:** The primary outcome measure was self-reported hearing aid benefit, measured using the Abbreviated Profile of Hearing Aid Benefit (APHAB). Secondary measures included the International Outcome Inventory for Hearing Aids (IOI-HA) and speech recognition in noise measured using the QuickSIN and digits-in-noise (DIN) tests. All measures were completed at baseline and at two intervals following hearing aid fitting (two- and six weeks).

**Results:** Sixty-four participants were included in the analytic sample, with equal number of participants (*n* = 32) randomized into SF and AF groups. The groups did not differ significantly in age (effect size *r*, -0.2; 95 CI -0.3 to 0.2) or four-frequency pure tone average (PTA; effect size *r*, 0.2; 95% CI -0.1 to 0.4). After the two-week field trial, the SF group had an initial advantage compared to the AF group on the self-reported benefit measures (APHAB; Cohen ‘s *d*, -0.5; 95% CI -1.0 to 0, IOI-HA; effect size *r*, 0.3; 95% CI 0.0 to 0.5) but not speech recognition in noise. At the end of the six-week trial, no meaningful differences were evident between the groups on any outcome measures.

**Conclusion and relevance:** This effectiveness trial indicates self-fitting OTC hearing aids with remote support provides comparable outcomes to a hearing aid fitted using audiologist best practices at 6-weeks post-fitting. Self-fitting OTC hearing aids can provide an effective intervention for mild to moderate hearing loss.

**KEY POINTS:** *Question:* Can self-fitting over-the-counter (OTC) hearing aids provide similar outcomes compared to best-practice audiologist-fitted hearing aids?

*Findings:* Self-reported and speech-in-noise benefit was equivalent between the self-fitting OTC and audiologist-fitted hearing aid conditions at the end of the six-week trial.

*Meaning:* A self-fitting OTC hearing aid can be an effective intervention option for people with mild-to-moderate hearing loss and produce self-perceived and clinical outcomes similar to that of a clinical audiologist-fitted hearing aid.

## INTRODUCTION

A person with hearing loss can benefit from a range of interventions to curtail detriment to quality of life. Most adults experience hearing loss which is sensorineural and permanent, leaving hearing aids as the most common intervention option ^1^. Unfortunately, uptake and use of hearing aids is low even among populations with adequate access to audiological resources ^2-5^. In the United States, hearing aid use among adults who could benefit from them is estimated to be only 20% ^4^. The reasons for poor hearing aid adoption are varied, but major barriers have been access and affordability. Until recently, people with hearing loss could only obtain hearing aids after consultation with a credentialed dispenser.

A working group for accessible and affordable hearing care was established by the National Institute on Deafness and Communication Disorders. They identified priority research areas for progressing hearing care access, including the development of self-testing, self-fitting hearing aids ^6,7^. The President ‘s Council of Advisors on Science and Technology, and the National Academies of Sciences, Engineering and Medicine, both organizations that inform the American federal government, also highlighted the role that over-the-counter (OTC) hearing aids could play in addressing the accessibility gap ^8^. Consequently, the Food and Drug Administration (FDA) passed the Reauthorization Act of 2017 (FDARA), directing the creation of an OTC hearing aid category^9^. The final regulations were recently published and became effective from October 17, 2022 ^10^. This new category of preset OTC and self-fitting OTC hearing aids has quickly become available at a significantly reduced cost when compared to prescription hearing aids ^11^.

The concept of self-fitting hearing aids was introduced more than a decade ago ^12^. In summary, a self-fitting hearing aid has the following key properties: (i) an automated fitting or in-situ test approach, (ii) can be operated without a clinician, and (iii) provides the user options to alter settings using accompanying controls or software ^12,13^. Earlier research validated specific elements of the self-fitting process, including the accuracy of measuring pure-tone thresholds ^14-16^ and aspects of usability ^17,18^. Additionally, alternative procedures for gain prescription were validated, including using different preset fitting parameters ^19,20^ and user self-selected settings ^21^.

Preliminary data provided to FDA from various device manufacturers suggest OTC hearing aids can be an acceptable intervention for mild-to-moderate hearing loss. However, there are limited well-controlled clinical studies reporting on OTC efficacy and effectiveness, especially using devices on the market. The few published trials available support relatively equivalent performance outcomes ^19-21^, but none consider factors beyond the hearing aid like post-fitting support, troubleshooting or remote adjustments ^20^. This study compared the effectiveness of a self-fitting OTC hearing aid with remote support to an audiologist-fitted hearing aid using best-practice. The hypothesis was that self-reported outcomes and speech recognition in noise benefit of the self-fitting group would be non-inferior to the audiologist-fitted group.

## METHOD

### Study design

This randomized clinical trial was conducted at the University of Pretoria, South Africa, from April to August 2022. The Humanities Research Ethics Committee at the University of Pretoria reviewed and approved the study protocol, and all participants provided written informed consent before participation. The trial was registered at ClinicalTrials.gov (Identifier: NCT05337748). This parallel-designed study consisted of two arms, self-fitting (SF) and audiologist-fitted (AF), with equal participant allocation to the two groups. Due to the nature of the trial, blinding was not possible.

Three research audiologists, each registered with the Health Professions Council of South Africa, were involved in carrying out the procedures. SF participants were provided with Lexie Lumen self-fitting OTC hearing aids in their standard commercial packaging through the study audiologists ^22^. Lexie Lumen hearing aids are behind-the-ear digital hearing aids with 16 channels, wide-dynamic range compression, adaptive directionality, and noise reduction. As directed by the accompanying instructional materials (booklet), participants were required to download the Lexie smartphone app and follow the instructions to fit the hearing aids. The research audiologist provided no assistance or orientation. The self-fitting parameters were based on a proprietary algorithm and used in-situ threshold measurements (in frequencies 0.5, 1, 2, 3, 4, and 6 kHz) conducted through the hearing aids. Participants in the AF group were fitted with the same hearing aid by the research audiologists using hearing thresholds measured using a calibrated audiometer in a soundproof booth. Real-ear verification ensured that the hearing aid output matched a gold-standard fitting algorithm, i.e., National Acoustics Laboratories ‘ Non-Linear Version 2 (NAL-NL2) at 0.5, 1, 2, and 4 kHz within a 5 dB tolerance limit, considered best-practice clinical verification (see supplement 1) ^23-25^. Following this procedure, the audiologist instructed the participants on hearing aid use as done in routine clinical practice.

After the baseline evaluation, randomization was performed by the researcher using a random number generator. Hearing aid fitting was completed in a second session, and participants conducted a two-week field trial. During the first two weeks, no assistance or fine-tuning via the Lexie remote support service was allowed for the SF group, nor was adjustment by the audiologist for the AF group. This design procedure isolated the benefit of fitting from the influence of online support on the SF participants. At the first follow-up, the audiologist conducted fine-tuning for the AF group on request. SF participants were informed that they could contact remote support for troubleshooting or adjustment. After another four-week field trial, the final assessments were conducted.

### Outcome measures

The outcome measures were administered at three time points, including the baseline unaided condition (T0) and aided conditions at two-weeks (T1) and six-weeks post-fitting (T2). The Abbreviated Profile of Hearing Aid Benefit (APHAB) is a 24-item self-assessment inventory to rate communication difficulties in different listening situations ^26^, and was the primary outcome of the study. Ultimately, benefit is calculated as the comparison of unaided and aided scores. Four sub-scales are evaluated, namely (i) ease of communication, (ii) reverberation, (iii) background noise, and (iv) aversiveness. The global score is the average score for all subscales, excluding aversiveness. International Outcome Inventory for Hearing Aids (IOI-HA) ^27^ was also included as a secondary outcome measure for self-reported benefit and determined the effectiveness of a hearing aid intervention in seven domains. Benefit is rated using five ordinal response categories, from worst to best outcome. The IOI-HA was provided post-hearing aid fitting at the first follow-up (T1) and final assessment (T2).

Additional secondary outcomes were speech recognition in noise, evaluated using the QuickSIN ^28^ and digits-in-noise (DIN) ^29,30^ tests. The tests were conducted at baseline (unaided) and aided at the two follow-up sessions. The QuickSIN measures a signal-to-noise ratio (SNR) loss of hearing sentences presented in four-talker babble noise, consisting of 18 × 6 – sentence lists with one sentence per SNR level (+25 to 0 dB SNR). After conducting a practice list, participants were presented with three lists to obtain the QuickSIN score. Averaged scores of three lists are accurate to about 1.6 dB at the 95% confidence interval ^28^. Stimuli were presented at a comfortable listening intensity in the sound field at 0º azimuth with the listener seated 1 meter away from the loudspeaker. The DIN was performed in the same audiometric setup. The test measures the decibel (dB) SNR where a listener could accurately recognize 50% of 23 randomly presented digit triplets (e.g., 6-8-2) presented adaptively (one-up, one-down procedure) in speech-weighted background masking noise ^29^. Real-ear verification using live-speech mapping at 65dB SPL was used to determine and compare real-ear-aided responses between the two groups (supplement 1). Lastly, the number of participants requesting fine-tuning or support between the two groups was captured.

### Participants and eligibility criteria

Participants 18 years and older who self-reported mild to moderate hearing loss in response to an advertisement and had no history of outer or middle ear disease 90 days before study inquiry were invited to attend a baseline session to establish audiometric candidacy. Participants with normal hearing thresholds (< 20 dB HL) at all frequencies (0.5, 1, 2, and 4 kHz), had possible outer or middle ear pathology or, had severe hearing loss with thresholds at 0.5, 1, 2, and 4 kHz more than 80 dB HL at two or more frequencies, were excluded. As detailed in Figure 1, 68 participants met the inclusion criteria and provided written informed consent to participate, and 64 were used for analysis with equal allocation to each group. We recruited a sample size similar to Sabin et al. 2020 ^21^. Table 1 summarizes the sample characteristics of each group.

**Table 1.**
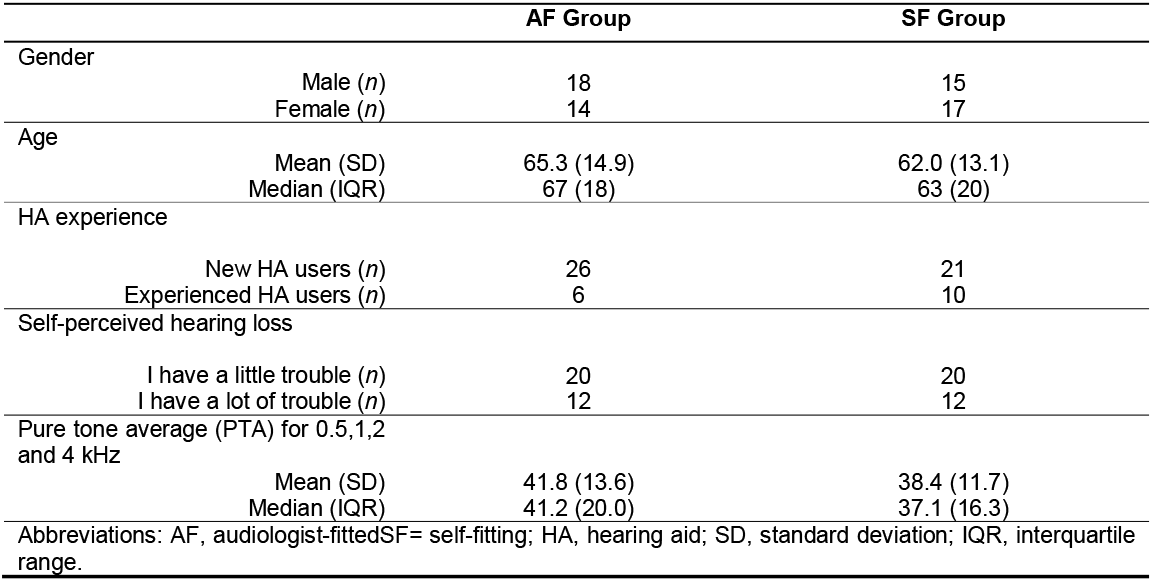
Summary characteristics of the AF and SF participants.

| **Table 1. Summary characteristics of the AF and SF participants** |  |  |  |
| --- | --- | --- | --- |
|  |  | **AF Group** | **SF Group** |
| Gender | Male (*n*) | 18 | 15 |
|  | Female (*n*) | 14 | 17 |
| Age | Mean (SD) | 65.3 (14.9) | 62.0 (13.1) |
|  | Median (IQR) | 67 (18) | 63 (20) |
| HA experience |  |  |  |
|  | New HA users (*n*) | 26 | 21 |
|  | Experienced HA users (*n*) | 6 | 10 |
| Self-perceived hearing loss |  |  |  |
|  | I have a little trouble (*n*) | 20 | 20 |
|  | I have a lot of trouble (*n*) | 12 | 12 |
| Pure tone average (PTA) for 0.5,1,2 and 4 kHz |  |  |  |
|  | Mean (SD) | 41.8 (13.6) | 38.4 (11.7) |
|  | Median (IQR) | 41.2 (20.0) | 37.1 (16.3) |
| Abbreviations: AF, audiologist-fitted; SF= self-fitting; HA, hearing aid; SD, standard deviation; IQR, interquartile range. |  |  |  |

**Figure 1.**
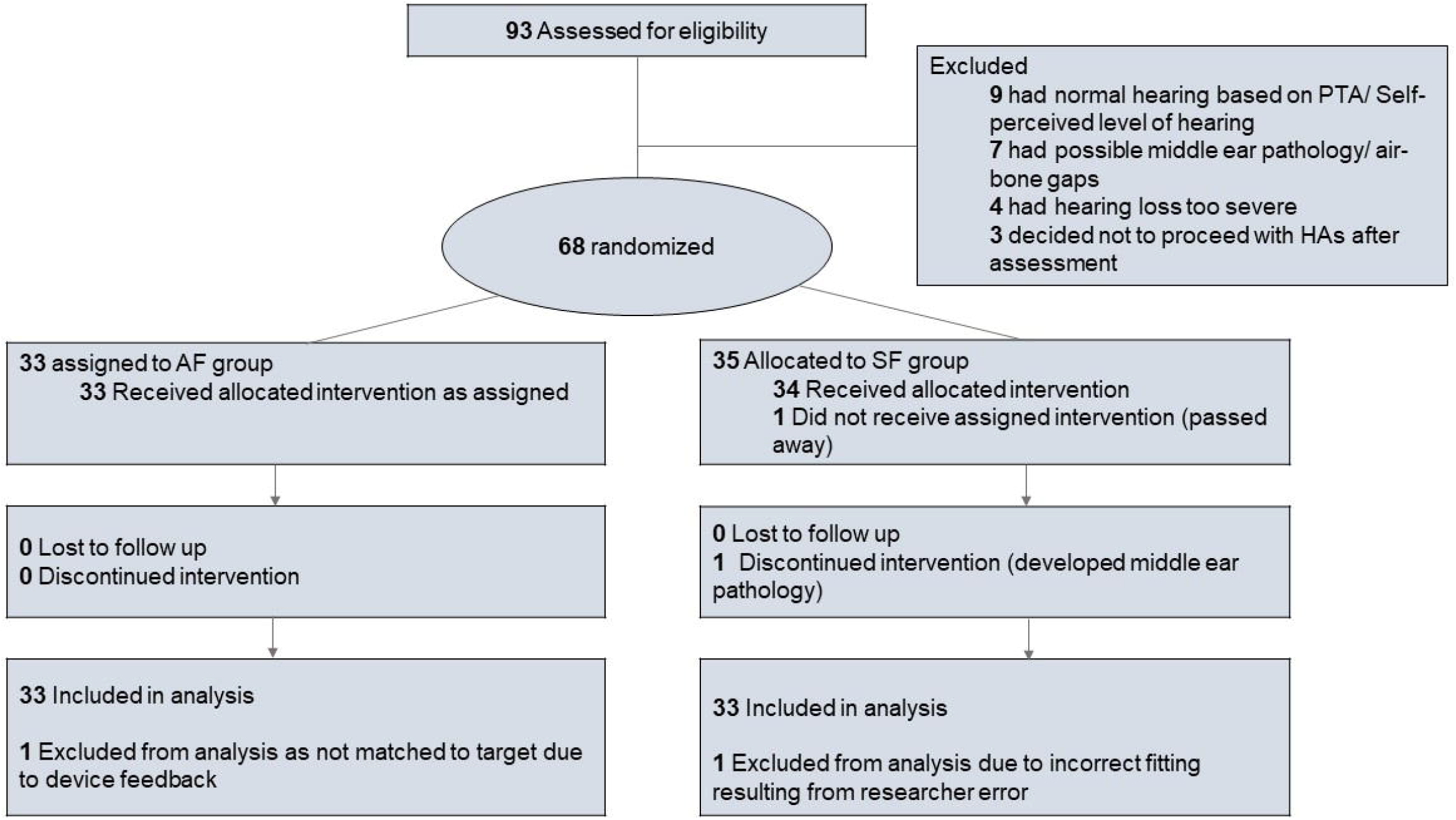
CONSORT flow diagram of the study. *AF, audiologist-fit group; SF, self-fitting.*

### Statistical analysis

We performed all statistical analyses using IBM SPSS statistics, version 28.0. Continuous variables (APHAB, QuickSIN, and DIN) were compared using Mann-Whitney U tests for non-normal distributed variables and independent samples *t*-tests for normal variables. IOI-HA scores were compared using the Mann-Whitney U test, as the results are ordinal response categories. Clinically meaningful differences were reported considering effect size and 95% CI. Cohen *d* was used for *t*-test, and effect size *r* calculated as z√N for Mann-Whitney U tests. Cohen *d* effect size was interpreted as small (*d* ≤ 0.2), small to medium (0.2 < *d* <0.5), medium (*d* = 0.5), medium to large (0.5 < *d* < 0.8), and large (*d* ≥ 0.8) ^31^ and effect size for Mann Whitney U tests as small (*r* ≤ 0.1), small to medium (0.1 < *r* < 0.30), medium (*r* = 0.3), medium to large (0.3 < *r* < 0.5) and large (*r* ≥ 0.5) ^31^. Effect sizes were considered clinically meaningful when they were medium or larger.

## RESULTS

### Self-reported outcomes

The two groups did not differ significantly in age (effect size *r*, -0.2; 95 CI -0.3 to 0.2) or PTA (effect size *r*, 0.2; 95% CI -0.1 to 0.4) (Table 1; supplement 2). Furthermore, the participants were fairly balanced in terms of gender (56% and 47% males in the AF and SF groups) and proportion of those with prior hearing aid experience (19% and 31% for the AF and SF groups).

Unaided baseline scores across all subscales and the global score of the APHAB were not significantly different between the groups (Table 2). Two weeks-post fitting (T1), the SF group achieved meaningfully better performance in background noise (mean difference, 10.1; Cohen *d*, 0.6; 95% CI 0.1 to 1.1) and on the APHAB global benefit scores (mean difference, 6.6; Cohen *d*, -0.5; 95% CI -1.0 to 0; Figure 2). After the six-week field trial (T2), the differences were not meaningful on any subscale or global benefit scores between the groups (Table 2; Figure 2). However, at T2 a higher proportion (87.5%) of SF participants scored above the 90% critical difference (9.9) for the APHAB ^32^ compared to the AB group (65.5%), although the overall effect size was small.

**Table 2.**
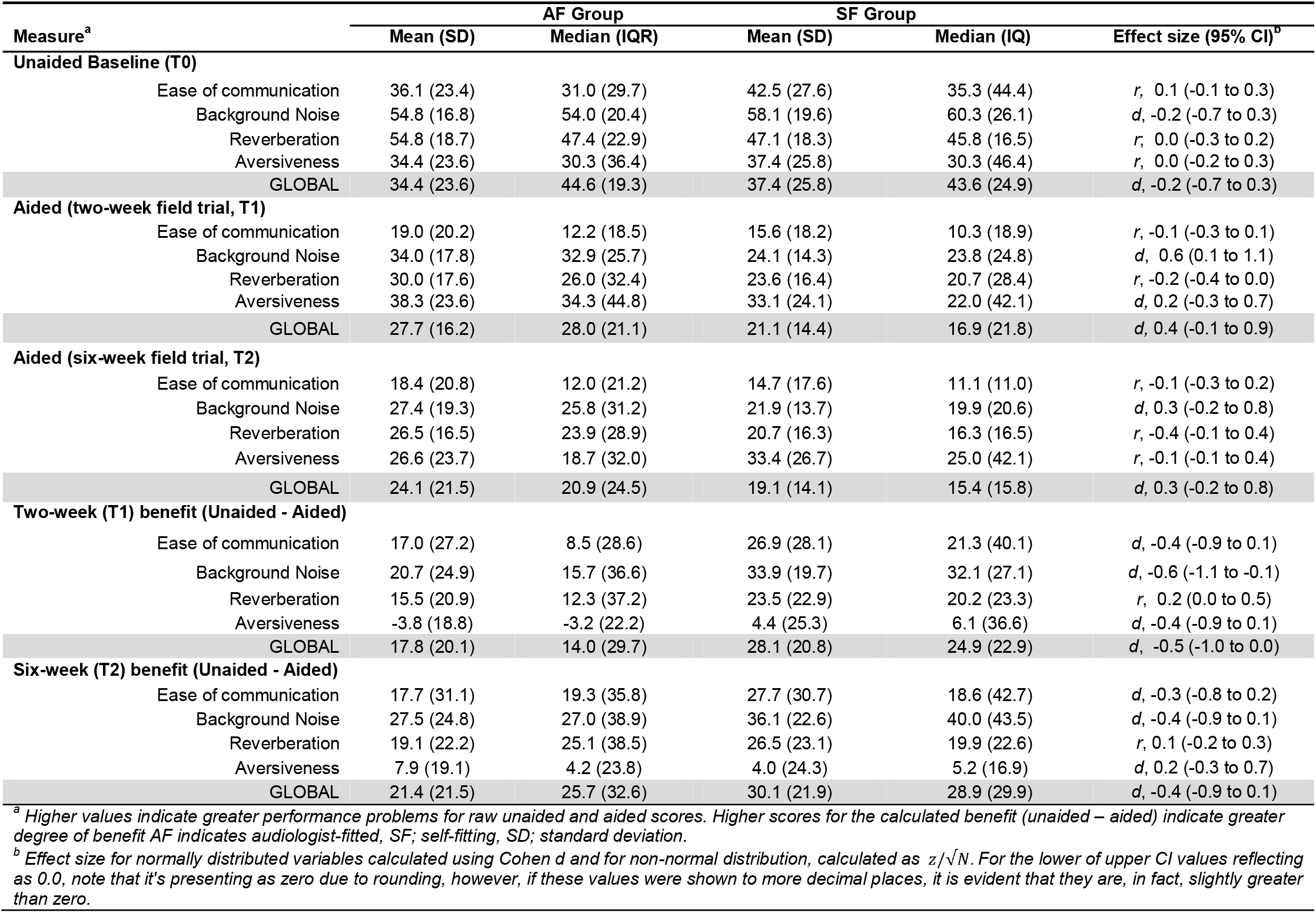
APHAB scores for the unaided baseline, follow-up, and benefit for the AF and SF group.

| Measure<sup>a</sup> | AF Group |  | SF Group |  | Effect size (95% CI)<sup>b</sup> |
| --- | --- | --- | --- | --- | --- |
|  | Mean (SD) | Median (IQR) | Mean (SD) | Median (IQ) |  |
| Unaided Baseline (T0) |  |  |  |  |  |
| Ease of communication | 36.1 (23.4) | 31.0 (29.7) | 42.5 (27.6) | 35.3 (44.4) | *r*, 0.1 (-0.1 to 0.3) |
| Background Noise | 54.8 (16.8) | 54.0 (20.4) | 58.1 (19.6) | 60.3 (26.1) | *d*, -0.2 (-0.7 to 0.3) |
| Reverberation | 54.8 (18.7) | 47.4 (22.9) | 47.1 (18.3) | 45.8 (16.5) | *r*, 0.0 (-0.3 to 0.2) |
| Aversiveness | 34.4 (23.6) | 30.3 (36.4) | 37.4 (25.8) | 30.3 (46.4) | *r*, 0.0 (-0.2 to 0.3) |
| GLOBAL | 34.4 (23.6) | 44.6 (19.3) | 37.4 (25.8) | 43.6 (24.9) | *d*, -0.2 (-0.7 to 0.3) |
| Aided (two-week field trial, T1) |  |  |  |  |  |
| Ease of communication | 19.0 (20.2) | 12.2 (18.5) | 15.6 (18.2) | 10.3 (18.9) | *r*, -0.1 (-0.3 to 0.1) |
| Background Noise | 34.0 (17.8) | 32.9 (25.7) | 24.1 (14.3) | 23.8 (24.8) | *d*, 0.6 (0.1 to 1.1) |
| Reverberation | 30.0 (17.6) | 26.0 (32.4) | 23.6 (16.4) | 20.7 (28.4) | *r*, -0.2 (-0.4 to 0.0) |
| Aversiveness | 38.3 (23.6) | 34.3 (44.8) | 33.1 (24.1) | 22.0 (42.1) | *d*, 0.2 (-0.3 to 0.7) |
| GLOBAL | 27.7 (16.2) | 28.0 (21.1) | 21.1 (14.4) | 16.9 (21.8) | *d*, 0.4 (-0.1 to 0.9) |
| Aided (six-week field trial, T2) |  |  |  |  |  |
| Ease of communication | 18.4 (20.8) | 12.0 (21.2) | 14.7 (17.6) | 11.1 (11.0) | *r*, -0.1 (-0.3 to 0.2) |
| Background Noise | 27.4 (19.3) | 25.8 (31.2) | 21.9 (13.7) | 19.9 (20.6) | *d*, 0.3 (-0.2 to 0.8) |
| Reverberation | 26.5 (16.5) | 23.9 (28.9) | 20.7 (16.3) | 16.3 (16.5) | *r*, -0.4 (-0.1 to 0.4) |
| Aversiveness | 26.6 (23.7) | 18.7 (32.0) | 33.4 (26.7) | 25.0 (42.1) | *r*, -0.1 (-0.1 to 0.4) |
| GLOBAL | 24.1 (21.5) | 20.9 (24.5) | 19.1 (14.1) | 15.4 (15.8) | *d*, 0.3 (-0.2 to 0.8) |
| Two-week (T1) benefit (Unaided - Aided) |  |  |  |  |  |
| Ease of communication | 17.0 (27.2) | 8.5 (28.6) | 26.9 (28.1) | 21.3 (40.1) | *d*, -0.4 (-0.9 to 0.1) |
| Background Noise | 20.7 (24.9) | 15.7 (36.6) | 33.9 (19.7) | 32.1 (27.1) | *d*, -0.6 (-1.1 to -0.1) |
| Reverberation | 15.5 (20.9) | 12.3 (37.2) | 23.5 (22.9) | 20.2 (23.3) | *r*, 0.2 (0.0 to 0.5) |
| Aversiveness | -3.8 (18.8) | -3.2 (22.2) | 4.4 (25.3) | 6.1 (36.6) | *d*, -0.4 (-0.9 to 0.1) |
| GLOBAL | 17.8 (20.1) | 14.0 (29.7) | 28.1 (20.8) | 24.9 (22.9) | *d*, -0.5 (-1.0 to 0.0) |
| Six-week (T2) benefit (Unaided - Aided) |  |  |  |  |  |
| Ease of communication | 17.7 (31.1) | 19.3 (35.8) | 27.7 (30.7) | 18.6 (42.7) | *d*, -0.3 (-0.8 to 0.2) |
| Background Noise | 27.5 (24.8) | 27.0 (38.9) | 36.1 (22.6) | 40.0 (43.5) | *d*, -0.4 (-0.9 to 0.1) |
| Reverberation | 19.1 (22.2) | 25.1 (38.5) | 26.5 (23.1) | 19.9 (22.6) | *r*, 0.1 (-0.2 to 0.3) |
| Aversiveness | 7.9 (19.1) | 4.2 (23.8) | 4.0 (24.3) | 5.2 (16.9) | *d*, 0.2 (-0.3 to 0.7) |
| GLOBAL | 21.4 (21.5) | 25.7 (32.6) | 30.1 (21.9) | 28.9 (29.9) | *d*, -0.4 (-0.9 to 0.1) |

<sup>a</sup> Higher values indicate greater performance problems for raw unaided and aided scores. Higher scores for the calculated benefit (unaided – aided) indicate greater degree of benefit AF indicates audiologist-fitted, SF; self-fitting, SD; standard deviation.
<sup>b</sup> Effect size for normally distributed variables calculated using Cohen *d* and for non-normal distribution, calculated as $z/\sqrt{N}$ . For the lower of upper CI values reflecting as 0.0, note that it's presenting as zero due to rounding, however, if these values were shown to more decimal places, it is evident that they are, in fact, slightly greater than zero.

**Figure 2.**
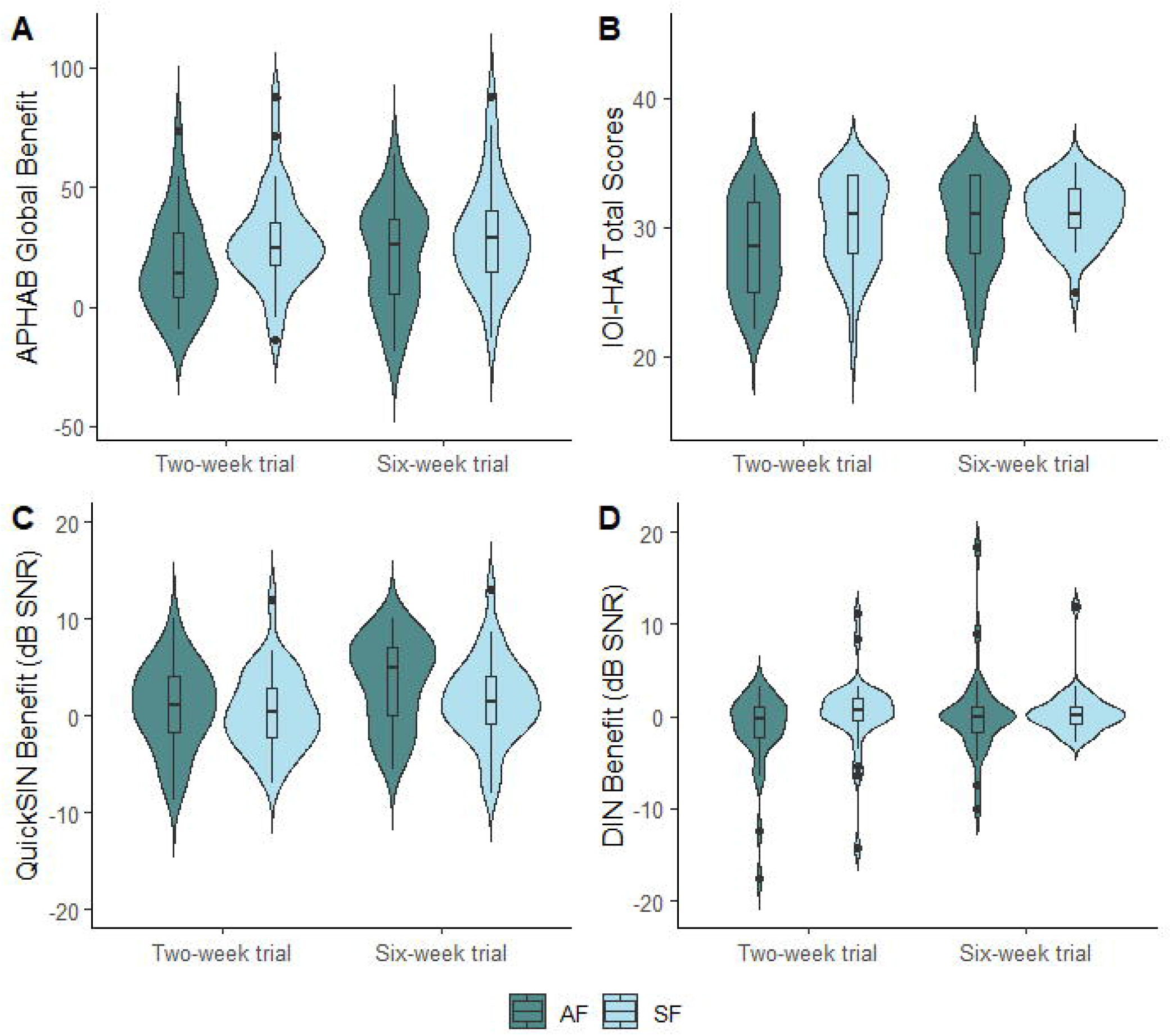
Outcome measures across the trial between the AF and SF group measured at two-weeks (T1) and six-weeks (T2) post hearing aid fitting. (A) APHAB global benefit scores, (B) IOI-HA total scores, (C) QuickSIN benefit scores, and (D) DIN benefit scores. Violin plots indicate kernel probability density. Boxes are interquartile range with median, and whiskers are 1.5 times the interquartile range. *AF indicates audiologist-fit; SF, self-fit.*

Based on the IOI-HA conducted at T1, SF participants reported meaningfully longer hearing aid use per day (effect size *r* = 0.31; 95% CI) (Table 3). None of the other individual items were significantly different between the groups. However, the total score was meaningfully better for the SF compared to AF group (effect size *r =* 0.30; 95% CI 0.0 to 0.5) at T1. After the six-week trial (T2), none of the IOI-HA items or total score were significantly different between the groups.

**Table 3.** IOI-HA scores for the AF and SF group after a two and six-week field trial.

| IOI-HA Subscales and score | AF Group |  | SF Group |  | Effect size (95% CI)<sup>a</sup> |
| --- | --- | --- | --- | --- | --- |
|  | Mean (SD) | Median (IQR) | Mean (SD) | Median (IQR) |  |
| **Two-week trial (T1)** |  |  |  |  |  |
| Q1 - Use | 3.9 (0.8) | 4.0 (0.8) | 4.4 (0.7) | 4.0 (1.0) | *r*, 0.3 (0.1 to 0.6) |
| Q2 - Benefit | 3.9 (1.0) | 4.0 (2.0) | 4.3 (0.7) | 4.0 (1.0) | *r*, 0.2 (0.0 to 0.5) |
| Q3 – Residual activity limitations | 4.0 (0.8) | 4.0 (2.0) | 4.3 (0.6) | 4.0 (1.0) | *r*, 0.2 (0.0 to 0.5) |
| Q4 - Satisfaction | 4.4 (0.7) | 5.0 (1.0) | 4.3 (1.1) | 5.0 (1.8) | *r*, 0.0 (-0.2 to 0.3) |
| Q5 – Residual participation restrictions | 3.8 (1.1) | 4.0 (2.0) | 4.0 (1.3) | 5.0 (1.0) | *r*, 0.2 (-0.1 to 0.4) |
| Q6 – Impact on others | 4.1 (1.3) | 5.0 (2.0) | 4.5 (1.0) | 5.0 (1.0) | *r*, 0.2 (-0.1 to 0.4) |
| Q7 – Quality of life | 4.2 (0.9) | 4.0 (1.8) | 4.3 (0.7) | 4.0 (1.0) | *r*, 0.1 (-0.2 to 0.3) |
| Total Score<sup>b</sup> | 28.4 (3.7) | 28.5 (7.0) | 30.3 (3.4) | 31.0 (6.0) | *r*, 0.3 (0.0 to 0.5) |
| **Six-week trial (T2)** |  |  |  |  |  |
| Q1 - Use | 4.1 (0.7) | 4.0 (1.0) | 4.4 (.6) | 4.0 (1.0) | *r*, 0.2 (-0.1 to 0.4) |
| Q2 - Benefit | 4.3 (0.9) | 4.5 (1.0) | 4.6 (.8) | 5.0 (1.0) | *r*, 0.1 (-0.1 to 0.4) |
| Q3 – Residual activity limitations | 4.2 (0.7) | 4.0 (1.0) | 4.3 (.6) | 4.0 (1.0) | *r*, 0.1 (-0.2 to 0.3) |
| Q4 - Satisfaction | 4.6 (0.5) | 5.0 (1.0) | 4.8 (.6) | 5.0 (0) | *r*, 0.2 (-0.1 to 0.4) |
| Q5 – Residual participation restrictions | 4.2 (1.1) | 5.0 (1.0) | 4.3 (1.2) | 4.0 (1.0) | *r*, -0.1 (-0.3 to 0.1) |
| Q6 – Impact on others | 4.3 (1.2) | 5.0 (1.0) | 4.5 (1.1) | 5.0 (1.0) | *r*, 0.2 (-0.1 to 0.4) |
| Q7 – Quality of life | 4.4 (.8) | 5.0 (1.0) | 4.5 (1.1) | 5.0 (1.0) | *r*, 0.0 (-0.2 to 0.3) |
| Total Score<sup>b</sup> | 30.2 (3.5) | 31.0 (6.0) | 30.7 (3.1) | 31.0 (3.0) | *r*, 0.1 (-0.1 to 0.4) |

<sup>a</sup>Calculated as $z/\sqrt{n}$ . For the lower of upper CI values reflecting as 0.0, note that it's presenting as zero due to rounding, however, if these values were shown to more decimal places, it is evident that they are, in fact, slightly greater than zero.
<sup>b</sup>Calculated as the sum of all seven IOI-HA items
IOI-HA indicates *International Outcomes Inventory for Hearing aids*, AF; *audiologist-fitted* and SF; *self-fitting*.

### Speech-in-noise outcomes

At baseline (T0), there were no significant differences between the groups for either the QuickSIN (Cohen *d*, 0.3; 95% CI 0.8 to -0.2) or DIN test (effect size *r*, -0.1; 95% CI -0.3 to 0.2). Benefit scores were determined by subtracting aided from unaided scores (Figure 2C & D). DIN benefit scores were not meaningfully different between the groups at either two weeks (effect size *r*, 0.2; 95% CI -0.2 to 0.3) or six weeks (effect size *r*, 0.1; 95% CI -0.2 to 0.3. Consistent with the DIN results, QuickSIN benefit was also not meaningfully different between the groups at two weeks (Cohen *d,* 0.1; 95% CI -0.4 to 0.6) or six weeks (Cohen *d,* 0.4; 95% CI -0.1 to 0.9). However, at six-weeks (T2), the proportion of participants performing better than the 90% critical difference for the QuickSIN (1.8 dB) ^28^ was 62.5% and 43.8% for the AF and SF group, respectively.

### Support requested

Frequency of requested hearing aid adjustment and support for the groups between two- and six-week post-fitting is worth noting. At the two-week aided assessment (T1), 65.6% of the AF participants requested fine-tuning compared to 6.3% of SF participants requesting remote support from the call center.

### Adverse events

Two days following hearing aid fitting, one participant in the SF group developed middle ear infection, a medical contraindication for using a self-fitting OTC or audiologist-fitted hearing aid. The participant was asked to discontinue intervention. No other adverse events were observed during the six-week trial.

## DISCUSSION

In this parallel design randomized effectiveness trial, the SF group performed comparably to the AF group. At two-weeks, the SF group had a small but meaningful advantage on two of the four outcome measures. After support and fine-tuning were provided to the SF (remote support) and AF groups, no clinically meaningful differences were evident in any outcome measures at the end of the six-week trial.

Audiologists fitted the AF group by matching the hearing aid gain according to prescriptive targets based on the audiometric results. Counseling and hearing aid orientation was provided in person at the fitting stage (T0), and hearing aids were fine-tuned based on the patient report at the two-week (T1) follow-up appointment. We considered this characteristic of clinical best practice, aligned with the American Speech-Language and Hearing Association guidelines ^33^. If this clinical model is the standard to which self-fitting OTC hearing aids are to be held, a key matter is whether the outcomes are similar between these two models. Self-reported hearing aid outcomes are standard outcome measures in hearing aid trials, especially since they strongly predict consistent hearing aid use ^34^.

We found better self-reported outcomes for the SF compared to the AF group after two-weeks of field use that was clinically meaningful. Specifically, the APHAB self-reported background noise performance was better for the SF group, as was the global benefit score. The SF group also showed a longer duration of daily hearing aid use on the IOI-HA, along with the total score. However, after six weeks of hearing aid use, the self-reported outcomes were not meaningfully different between the groups. Three previous effectiveness trials showed similar findings. The study by Humes and colleagues in 2017 used an alternative OTC method where users selected from different pre-programmed hearing aids, compared to an audiologist-fitted hearing aid and placebo control ^19^. Two self-reported outcome measures (Profile of Hearing Aid Benefit and the Hearing Handicap Inventory for the Elderly) were equivalent between the pre-programmed OTC and audiologist-fit hearing aids. However, a slight non-significant advantage was found for the audiologist-fitted group. These findings were replicated in 2019 using less restrictive participant selection ^20^. Sabin et al. [21] compared an audiologist best practice group to a self-fitting group fitted with the Bose Sound Control™ hearing aid. Here, users could select their own fitting parameters, including gain and spectral tilt. APHAB global scores and the short form of the Speech Spatial and Qualities of Hearing Scale (SSQ-12) showed no significant differences between the groups. The latter study is closely aligned to the current study through inclusion of a OTC hearing aid currently in the market. The weight of the evidence thus far supports self-reported benefits for a self-fitting OTC model to be comparable to an audiologist-fitted model for mild to moderate hearing loss.

There is a prevailing assumption that hearing aids matched to prescriptive targets through probe tube verification results in better speech recognition in noise ^35^. However, a recent systematic review indicates that although real-ear verified hearing aids are positively related to better speech recognition in noise, the pooled effect sizes are small ^35^, and the absolute dB SNR benefit is slight and clinically not meaningful ^36^. Our study showed little differences in speech in noise performance (i.e., QuickSIN and DIN scores) between the AF group matched to the prescriptive target and the SF groups at two- and six-weeks post-hearing aid fitting. Similarly, Sabin et al. [21] found no significant differences in speech recognition benefit (QuickSIN) between the audiologist best practice and SF groups when fitted with the same hearing aid ^21^. Considering the results of this and other studies, the current study suggests that target-matched hearing aids are not likely to produce functionally different outcomes in speech recognition in noise when compared to good self-fitting algorithms. Furthermore, observations presented here (supplement 1) and elsewhere suggest that matching gain to a prescriptive target may not necessarily be a comfortable starting point. For example, Sabin et al. [21] showed that participants preferred their self-selected fitting parameters, which were generally lower than the audiologist-selected gain ^21^. A few other studies on self-fitting hearing aids have similarly shown a preference for lower gain, especially in the higher frequency region ^25,37,38^. The OTC hearing aid in this study also applied lower gain in high-frequencies (supplement 1), which could, at least in part, contribute to the initial superior APHAB and IOI-HA benefit scores. In this study, the initial fitting for the AF group was to be matched as closely as possible to the prescriptive target. In a typical clinical setting, the audiologist would normally make adjustments to the fitting based on patient report generally lowering the hearing aid gain. Since the AF group more frequently requested fine-tuning than the SF group during the last four weeks, the hearing aid adjustments in the AF group likely contributed to more uniform outcomes at the end of the six-week trial.

The study had a few limitations. First, blinding was not possible. Furthermore, we investigated only one self-fitting OTC device with one fitting method. It is possible that other devices and fitting methods may produce outcomes with variable success. Finally, the results only report outcomes six-weeks post-fitting. Further field research investigating long-term outcomes is needed.

## CONCLUSIONS

This study demonstrates that the short-term outcomes of self-fitting OTC hearing aids for people with mild to moderate hearing loss are comparable to those obtained from audiologist-fitted hearing aids using best practices. Affordable self-fitting OTC hearing aids can be an accessible hearing intervention option with outcomes similar to the same audiologist-fit hearing aid.

## Data Availability

All data produced in the present study are available upon reasonable request to the authors

## CONFLICT OF INTEREST DISCLOSURE

Dr De Sousa has a relationship with the hearX group which includes consulting. Dr Swanepoel and Dr Moore have a relationship with the hearX Group (Pty) Ltd, which includes equity, consulting and potential royalties.

## FUNDING SUPPORT

This study received funding from the hearX (Pty) Ltd Group and National institutes of Health (1R21DC019598). R21/R33 Mobile technologies for delivering hearing care through community health workers. Primary investigators: D Swanepoel (UP) and David Moore (CCHM).

## ROLE OF THE FUNDER/ SPONSOR

The funder provided the Lexie Lumen devices and software support to complete data collection.

## Notes

### Clinical Trial

NCT05337748

### Author Declarations

The Humanities Research Ethics Committee at the University of Pretoria reviewed and approved the study protocol, and all participants provided written informed consent before participation. The trial was registered at ClinicalTrials.gov (Identifier: NCT05337748).

