## Supplement 1 for "Effectiveness of an over-the-counter self-fitting hearing aid compared to an audiologist-fitted hearing aid: A randomized clinical trial"

**SUPPLEMENTARY MATERIAL 1**

*Probe tube verification using speech mapping for the self-fit (SF) versus audiologist-fit (AF) group*

The figure below provides the comparison of the real-ear output (65 dB SPL) measured for the AF and SF group against NAL-NL2 prescriptive target conducted at the time of hearing aid fitting (Figure A and C). The groups were balanced with 32 participants in each group. Overall, a good match to target was obtained for the AF group (Figure 5A), with real-ear output measured at 0.25 to 6 kHz within a ± 5 dB tolerance limit across all frequencies, considered best-practice clinical verification.

Following the two-week field trial (T1), participants continued to wear the same hearing aids for approximately four additional weeks.  Fine tuning was conducted by the audiologist for 21/32 (65.6%) of the participants in the AF group. Participants in the SF group were informed that they could request remote support and fine-tuning by contacting the Lexie online call center, of which only two participants made use of the service. Real ear output after fine-tuning conducted at T1 for the AF and SF participants are presented in Figure B and D below.

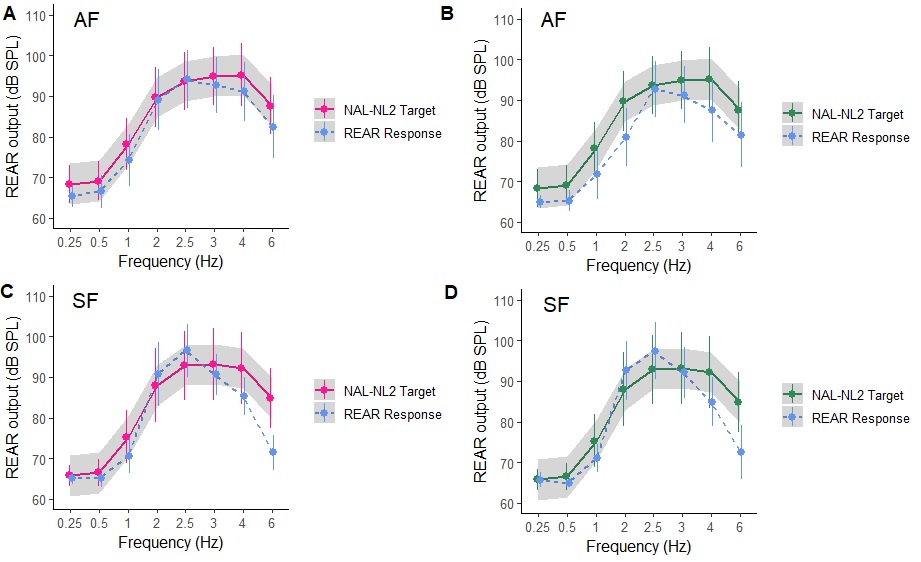

*Figure.* Comparison of prescribed NAL-NL2 real-ear targets and measured real-ear levels in dB SPL (blue) for left and right ears combined. (A) Comparison of NAL-NL2 target and real ear output in dB SPL for the AF group at initial fitting. (B) Comparison of NAL-NL2 target and real ear output in dB SPL for the AF group at 6 weeks, (C)Comparison of the NAL-NL2 target and real ear output in dB SPL for the SF group at fitting, (D) Comparison of the NAL-NL2 target and real ear output in dB SPL at 6 weeks. The stimulus was a 65 dB SPL speech signal (International Speech Test Signal) for the speech-mapping features for the MedRx system. Symbol = mean, error bar = ±1 standard deviation. Shading indicates the tolerance limits (±5dB) from the mean for NAL-NL2 prescriptive targets.
