## Supplement 2 for "Effectiveness of an over-the-counter self-fitting hearing aid compared to an audiologist-fitted hearing aid: A randomized clinical trial"

**SUPPLEMENTARY MATERIAL 2**

*Pure tone audiometric results for the self-fit OTC (SF) versus audiologist-fit (AF) group.*

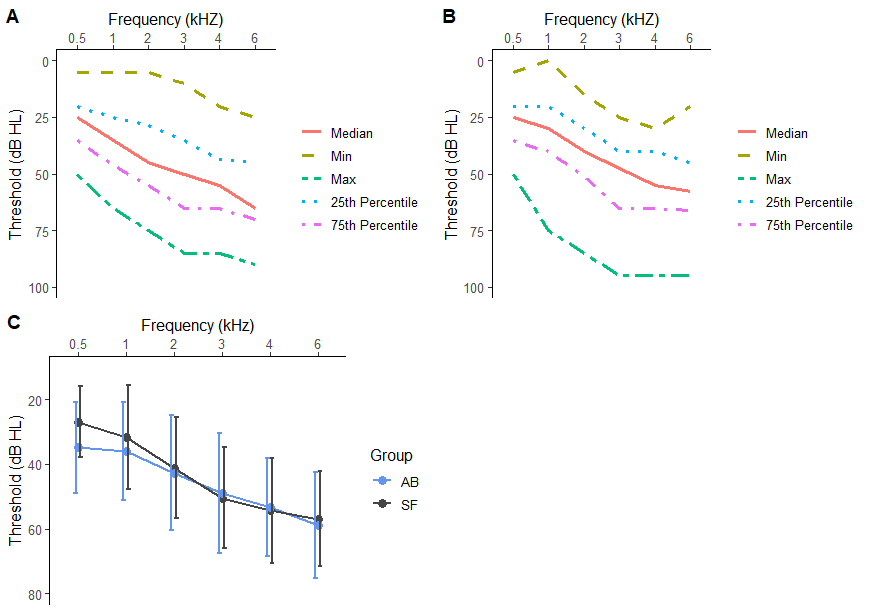

***Figure.*** Distribution of conventional pure tone audiometric frequencies combined left and right. (A) Distribution of pure tone thresholds for AB group, (B) Distribution of pure tone thresholds for the SF group, (C) Mean thresholds at 0.5 to 6 kHz, with error bars as standard deviation for the SF and AB groups. Abbreviations. SF, self-fit; AB, audiologist best practice.
